# Seek COVER: Development and validation of a personalized risk calculator for COVID-19 outcomes in an international network

**DOI:** 10.1101/2020.05.26.20112649

**Authors:** Ross D. Williams, Aniek F. Markus, Cynthia Yang, Talita Duarte Salles, Scott L. DuVall, Thomas Falconer, Jitendra Jonnagaddala, Chungsoo Kim, Yeunsook Rho, Andrew Williams, Amanda Alberga, Min Ho An, María Aragón, Carlos Areia, Edward Burn, Young Hwa Choi, Iannis Drakos, Maria Tereza Fernandes Abrahão, Sergio Fernández-Bertolín, George Hripcsak, Benjamin Skov Kaas-Hansen, Prasanna L Kandukuri, Jan A. Kors, Kristin Kostka, Siaw-Teng Liaw, Kristine E. Lynch, Gerardo Machnicki, Michael E. Matheny, Daniel Morales, Fredrik Nyberg, Rae Woong Park, Albert Prats-Uribe, Nicole Pratt, Gowtham Rao, Christian G. Reich, Marcela Rivera, Tom Seinen, Azza Shoaibi, Matthew E Spotnitz, Ewout W. Steyerberg, Marc A. Suchard, Seng Chan You, Lin Zhang, Lili Zhou, Patrick B. Ryan, Daniel Prieto-Alhambra, Jenna M. Reps, Peter R. Rijnbeek

**Author notes:** These authors contributed equally as co-first authors (Ross D. Williams, Aniek F. Markus). These authors contributed equally as co-last authors (Jenna M. Reps, Peter R. Rijnbeek). **Corresponding Author:** Peter R. Rijnbeek PhD, Department of Medical Informatics, Erasmus University Medical Center, Rotterdam, The Netherlands, Doctor Molewaterplein 403015 GD Rotterdam, The Netherlands.

## Abstract

**Objective:** To develop and externally validate COVID-19 Estimated Risk (COVER) scores that quantify a patient’s risk of hospital admission (COVER-H), requiring intensive services (COVER-I), or fatality (COVER-F) in the 30-days following COVID-19 diagnosis.

**Methods:** We analyzed a federated network of electronic medical records and administrative claims data from 14 data sources and 6 countries. We developed and validated 3 scores using 6,869,127 patients with a general practice, emergency room, or outpatient visit with diagnosed influenza or flu-like symptoms any time prior to 2020. The scores were validated on patients with confirmed or suspected COVID-19 diagnosis across five databases from South Korea, Spain and the United States. Outcomes included i) hospitalization with pneumonia, ii) hospitalization with pneumonia requiring intensive services or death iii) death in the 30 days after index date.

**Results:** Overall, 44,507 COVID-19 patients were included for model validation. We identified 7 predictors (history of cancer, chronic obstructive pulmonary disease, diabetes, heart disease, hypertension, hyperlipidemia, kidney disease) which combined with age and sex discriminated which patients would experience any of our three outcomes. The models achieved high performance in influenza. When transported to COVID-19 cohorts, the AUC ranges were, COVER-H: 0.69-0.81, COVER-I: 0.73-0.91, and COVER-F: 0.72-0.90. Calibration was overall acceptable.

**Conclusions:** A 9-predictor model performs well for COVID-19 patients for predicting hospitalization, intensive services and fatality. The models could aid in providing reassurance for low risk patients and shield high risk patients from COVID-19 during de-confinement to reduce the virus’ impact on morbidity and mortality.

## Introduction

The growing number of infections due to the Corona Virus Disease 2019 (COVID-19) has resulted in unprecedented pressure on healthcare systems worldwide, and a large number of casualties at a global scale. Although the majority of people have uncomplicated or mild illness (81%), some will develop severe disease leading to hospitalization and oxygen support (15%) or fatality (4%)^1,2^. The most common diagnosis in severe COVID-19 patients is pneumonia, other known complications include acute respiratory distress syndrome (ARDS), sepsis, or acute kidney injury (AKI)^1^. While there is currently no known cure or vaccine, the current approach to management of COVID-19 includes monitoring and controlling symptoms.

In response to the global pandemic, many countries have implemented measures aimed to reduce the average number of people a person with COVID-19 will infect^3-6^. The purpose of this was to prevent the spread of the virus, or at least to slow the spread, a process known as flattening the curve^7^. However, strategies such as social distancing have impacted economies globally and economic worries are causing countries to consider lifting measures earlier than epidemiologists recommend^8^. There are worries that this may cause a second wave of infections, as seen historically in other pandemics^9^. Multiple governments are starting to release de-confinement strategies, for example the state of New York^10^, British^11^, and Dutch^12^ governments have detailed plans to ease restrictions. However, they only concern population-level effects of likely disease spread and contain no information on how an individual’s risk impacts their likely morbidity and mortality if they were to contract the virus. Research has shown that COVID-19 does not impact all ages and sexes equally^13^ and as such a more personalised risk assessment can aid in improving outcomes. In a recent BMJ editorial^14^, the authors conclude that the COVID-19 response “is about protecting lives and communities most obviously at risk in our unequal society”. Quantifying a patient’s risk of having severe or critical illness when infected with COVID-19, could be used to help countries plan strategies to shield the most vulnerable patient populations. This is essential during the planning of de- confinement strategies.

The WHO Risk Communication Guidance distinguishes two categories of patients at high risk of severe disease: those older than 60 years and those with “underlying medical conditions” which is non-specific^15^. Using general criteria to assess the risk of poor outcomes is a crude risk discrimination mechanism as entire patient groupings are treated homogeneously ignoring individual differences. Prediction models can quantify a patient’s individual risk and data-driven methods could identify risk factors that have been previously overlooked. The number of studies developing prediction models for COVID-19 is still limited and of insufficient quality, as suggested in a recent systematic review ^16^. In a recent review the A-DROP model was recommend^17,18^, however this requires lab tests and thus requires a patient to be either in hospital or another setting to receive tests. Due to the high load on healthcare systems and the highly contagious nature of the disease, it is useful to have a model that can be used without this information. As such we propose three models that can assess based off historical and demographic information Previously published COVID-19 prediction models have been criticised for being i) poorly reported, ii) developed using small data samples, and iii) lacking external validation.

In this paper we aim to develop COVID-19 Estimated Risk (COVER) scores to quantify a patient’s risk of hospital admission (COVER-H), requiring intensive services (COVER-I), or fatality (COVER- F) due to COVID-19 using the Observational Health Data Sciences and Informatics (OHDSI) Patient-Level Prediction framework^19^. The research collaboration known as OHDSI has developed standards and tools that allow patient-level prediction models to be developed and externally validated rapidly following accepted best practices^20^. This allows us to overcome the previously identified shortcomings of previous COVID-19 prediction papers by reporting according to open science standards and implementing widespread external validation. To overcome the shortcoming of using small data for development, we made use of the abundant data from patients with influenza or flu-like symptoms to develop the models and then we tested whether the models transport to COVID-19 patients. Given the symptomatic similarities between the two diseases we hypothesized that the developed models will be able to transport between the two problem settings.

## Methods

We performed a retrospective cohort study to develop COVID-19 prediction models for severe and critical illness.

### Source of data

This study used observational healthcare databases from six different countries. All datasets used in this paper were mapped into the Observational Medical Outcomes Partnership Common Data Model (OMOP-CDM)^21^. The OMOP-CDM was developed for researchers to have diverse datasets in a consistent structure and vocabulary. This enables analysis code and software to be shared among researchers which facilitates external validation of the prediction models.

### Consent to publish

All databases obtained institutional review board (IRB) approval or used deidentified data that was considered exempt from IRB approval. Informed consent was not necessary at any site. The OMOP-CDM datasets used in this paper are listed in Table 1.

**Table 1.**
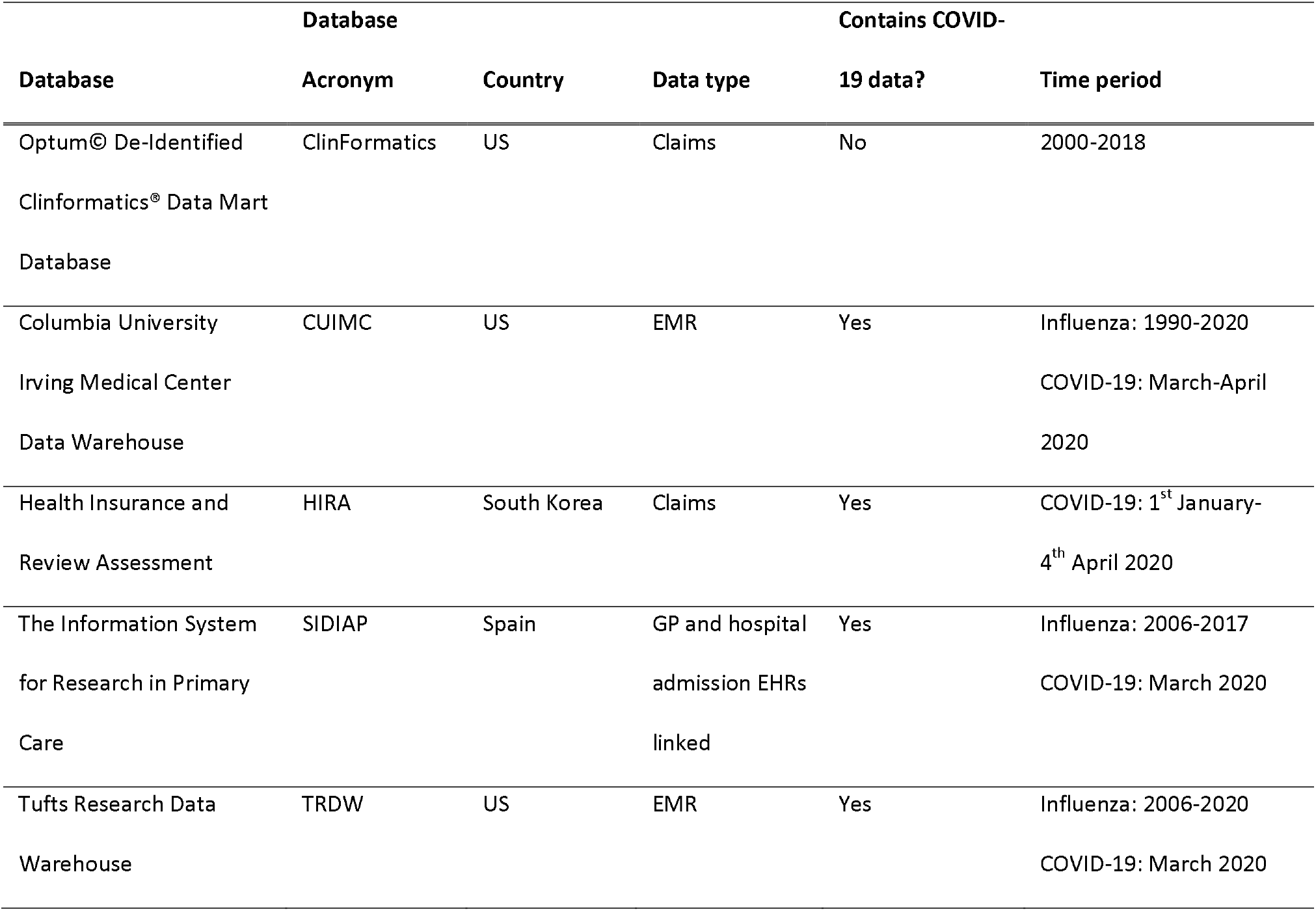
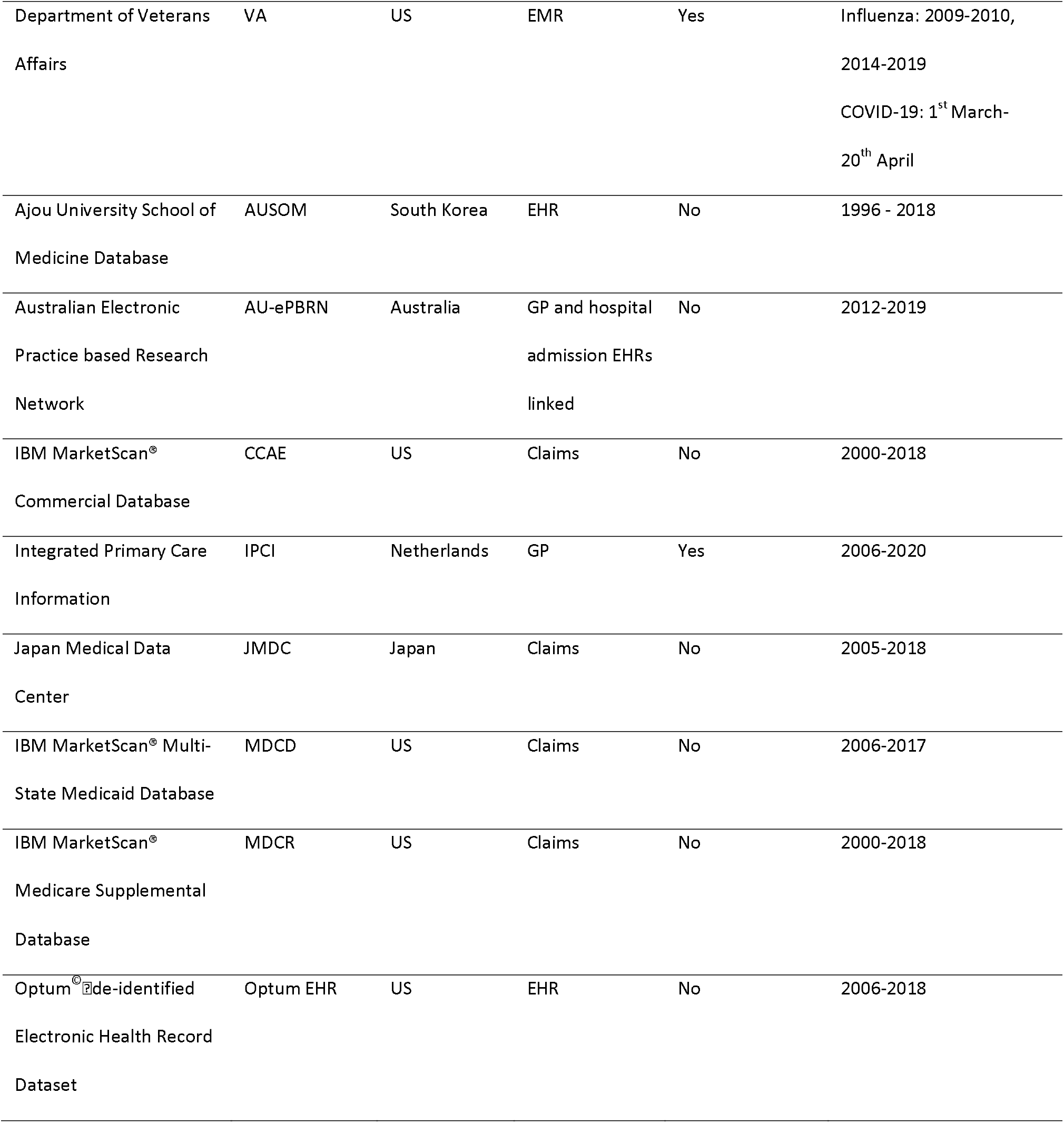
Data sources formatted to the Observational Medical Outcomes Partnership Common Data Model (OMOP-CDM) used in this research (data type: claims, electronic health/medical records (EHR/EMR), general practitioner (GP))

| Database |  |  |  | Contains COVID- |  |
| --- | --- | --- | --- | --- | --- |
| Database | Acronym | Country | Data type | 19 data? | Time period |
| Optum® De-Identified<br>Clinformatics® Data Mart<br>Database | ClinFormatics | US | Claims | No | 2000-2018 |
| Columbia University<br>Irving Medical Center<br>Data Warehouse | CUIMC | US | EMR | Yes | Influenza: 1990-2020<br>COVID-19: March-April<br>2020 |
| Health Insurance and<br>Review Assessment | HIRA | South Korea | Claims | Yes | COVID-19: 1<sup>st</sup> January-<br>4<sup>th</sup> April 2020 |
| The Information System<br>for Research in Primary<br>Care | SIDIAP | Spain | GP and hospital<br>admission EHRs<br>linked | Yes | Influenza: 2006-2017<br>COVID-19: March 2020 |
| Tufts Research Data<br>Warehouse | TRDW | US | EMR | Yes | Influenza: 2006-2020<br>COVID-19: March 2020 |
| Department of Veterans Affairs | VA | US | EMR | Yes | Influenza: 2009-2010,<br>2014-2019<br>COVID-19: 1<sup>st</sup> March-20<sup>th</sup> April |
| Ajou University School of Medicine Database | AUSOM | South Korea | EHR | No | 1996 - 2018 |
| Australian Electronic Practice based Research Network | AU-ePBRN | Australia | GP and hospital admission EHRs linked | No | 2012-2019 |
| IBM MarketScan® Commercial Database | CCAE | US | Claims | No | 2000-2018 |
| Integrated Primary Care Information | IPCI | Netherlands | GP | Yes | 2006-2020 |
| Japan Medical Data Center | JMDC | Japan | Claims | No | 2005-2018 |
| IBM MarketScan® Multi-State Medicaid Database | MDCD | US | Claims | No | 2006-2017 |
| IBM MarketScan® Medicare Supplemental Database | MDCR | US | Claims | No | 2000-2018 |
| Optum® de-identified Electronic Health Record Dataset | Optum EHR | US | EHR | No | 2006-2018 |

### Participants

For validation in COVID-19 we used a cohort of patients presenting at an initial healthcare provider interaction in a general practice (GP), emergency room (ER), or outpatient (OP) visit with COVID-19 disease. The initial healthcare provider interaction is used as index date, which is the point in time a patient enters a cohort. COVID-19 disease was identified by a diagnosis code for COVID-19 or a positive test for the SARS-COV-2 virus that was recorded after January 1^st^ 2020. We required patients to be aged 18 or over, have at least 365 days of observation time prior to the index date and no diagnosis of influenza, flu-like symptoms, or pneumonia in the preceding 60 days.

For model development, we identified patients aged 18 or over with a GP, ER, or OP visit with influenza or flu-like symptoms (e.g. fever and either cough, shortness of breath, myalgia, malaise, or fatigue), at least 365 days of prior observation, and no symptoms in the preceding 60 days.

### Outcome

We investigated three outcomes of COVID-19: 1) hospitalization with pneumonia from index up to 30 days after index, 2) hospitalization with pneumonia that required intensive services (ventilation, intubation, tracheotomy, or extracorporeal membrane oxygenation) or death after hospitalization with pneumonia from index up to 30 days after index, and 3) death from index up to 30 days after index.

The full details of the participant cohorts and outcomes used for development and validation can be found in the R packages.

### Predictors

When using a data-driven approach to model development, generally the resulting models contain a large number of predictors. We developed a data-driven model using age in groups (18-19, 20-25, 26-30, …, 95+), sex and binary variables indicating the presence or absence of recorded conditions and drugs any time prior to index. Missing records are thus effectively imputed as zero, exceptions are age and sex, which are always recorded in the OMOP-CDM. In total, we derived 31,917 candidate predictors indicating the presence of the 31,917 unique conditions/drugs recorded prior to the index date (GP, ER, or OP visit) for each patient. This may optimise performance, but a large number of predictors can be a barrier to clinical implementation. The utility of models for COVID-19 requires that they can be widely implemented across worldwide healthcare settings. Therefore, in addition to a data-driven model, we investigated two models that include fewer candidate predictors.

The age/sex model used age groups and sex as candidate predictors. The COVER scores included 7 candidate predictors, in addition to age groups and sex, that corresponded to the following conditions existing any time prior to the index date (GP, ER, or OP visit): cancer, chronic obstructive pulmonary disease, diabetes, heart disease, hypertension, hyperlipidemia and kidney disease (chronic and acute). Full details on how these 7 predictors were created as well as what constitutes a predictor can be found in Appendix 1A of the online supplement.

### Sample Size

The models were developed using the Optum^©^ De-Identified Clinformatics® Data Mart Database. We identified 7,344,117 valid visits with influenza or flu-like symptoms, of which 4,431,867 were for patients aged 18 or older, 2,977,969 of these had >= 365 days observation prior to the visit, and 2,082,277 of these had no prior influenza/symptoms/pneumonia in the 60 days prior to index. We selected a random sample of 150,000 patients from the total population to efficiently develop models to address the current pandemic, while preserving the outcome proportion. This allowed us to perform model development on a large dataset of flu patients whilst also leaving ∼2m patients for a validation to provide strong evidence of performance and reduce the probability the high performance achieved in the sample was due to over-fitting. Figure 1 is a flow chart demonstrating this.

**Fig 1.**
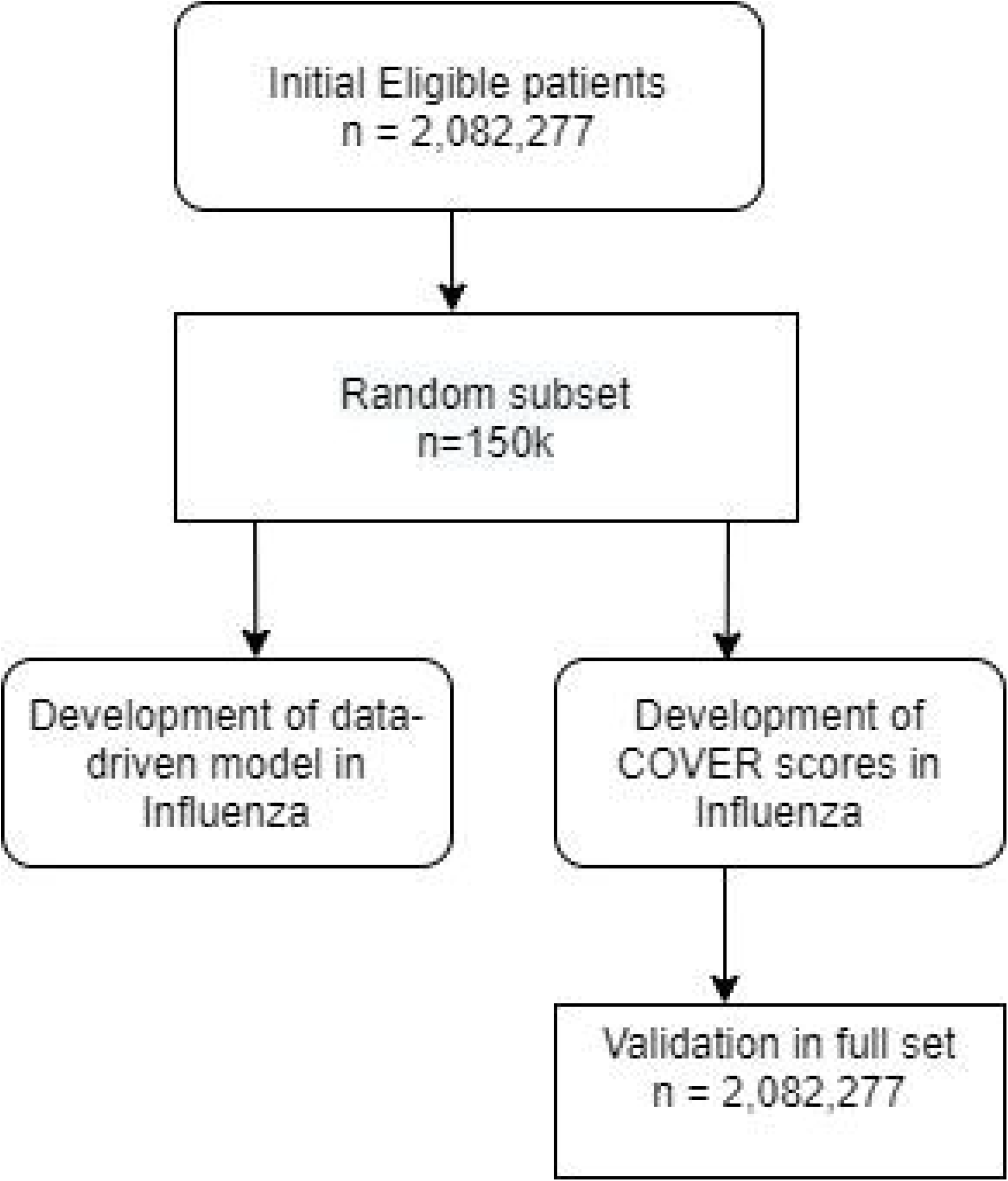

### Statistical analysis methods

Model development followed a previously validated and published framework for the creation and validation of patient-level prediction^19^. We used a person ‘train-test split’ method to perform internal validation. In each development cohort, a random split sample (‘training sample’) containing 75% of patients was used to develop the prediction models and the remaining 25% of patients (‘test sample’) was used to validate the models. We trained models using LASSO regularised logistic regression, using a 3-fold cross validation technique in the influenza training sample to learn the optimal regularization hyperparameter through an adaptive search^22^. We used R (version 3.6.3) and the OHDSI Patient-Level Prediction package (version 3.0.16) for all statistical analyses^19^.

To evaluate the performance, we calculate the overall discrimination of the model using the area under the receiver operating characteristic curve (AUC), the area under the precision recall-curve (AUPRC), and the model calibration. The AUC indicates the probability that for two randomly selected patients, the patient who gets the outcome will be assigned a higher risk. The AUPRC shows the trade-off between identifying all patients who get the outcome (recall) versus incorrectly identifying patients without outcome (precision) across different risk thresholds. The model calibration is presented in a plot to examine agreement between predicted and observed risks across deciles of predicted risk. Calibration assessment is then performed visually rather than using a statistic or numeric value as this provides a better impression of the direction and scale of miscalibration^23^. Summary statistics are reported from the test samples.

We performed two types of external validation. A classical external validation in which we applied the models to identical settings across diverse patient populations with influenza or flu-like symptoms prior to 2020 not used to develop the model, and a specific COVID-19 validation for databases containing COVID-19 data. To do this we assessed patients with confirmed COVID-19 in 2020. We examined the external validation using AUC, AUPRC and model calibration in the same way as internally.

This study was conducted and reported according to the Transparent Reporting of a multivariate prediction model for Individual Prediction or Diagnosis (TRIPOD) guidelines^24^ and adhered to the open science principles for publicly prespecifying and tracking changes to study objectives, protocol and code as described in the Book of OHDSI^25^. For transparency, the R packages for the development and external validation of the models in any database with OMOP-CDM are available on GitHub at: https://github.com/ohdsi-studies/Covid19PredictionStudies

## Results

### Online results

The complete results are available as an interactive app at: http://evidence.ohdsi.org/Covid19CoverPrediction

### Participants

Table 2 describes the characteristics at baseline of the patients across the different databases used for development and external validation. Out of the 150,000 patients sampled with influenza or flu-like symptoms in the development database (ClinFormatics), there were 6,712 patients requiring hospitalization with pneumonia, 1,828 patients requiring hospitalization and intensive services with pneumonia, and 748 patients died within 30 days. See Table 2 for the full outcome proportions across the databases included in this study. A total of 44,507 participants with COVID-19 disease were further included for external validation.

**Table 2.** Population size, outcome proportion and characteristics for the development database (influenza) and external validation in COVID-19 and influenza (N/A indicates this result is not available)

|  | Development | External validation: COVID-19 |  |  |  |  | External validation: influenza |  |  |  |  |  |  |  |
| --- | --- | --- | --- | --- | --- | --- | --- | --- | --- | --- | --- | --- | --- | --- |
|  | ClinFormati | CUI MC | HIRA | SIDIAP | TRDW | VA | AUSOM | AU-ePBRN | CCAE | IPCI | JMDC | MDCD | MDCR | Optum EHR |
| Number of participants | 2,082,277 | 2,731 | 1,985 | 37,950 | 395 | 1,446 | 3,105 | 2,791 | 3,146,801 | 29,132 | 1,276,478 | 536,806 | 248,989 | 1,654,157 |
| Hospitalization with pneumonia (Outcome proportion %) | 105,030<br>(5.04) | N/A | 89<br>(4.48) | 1,223<br>(1.11) | 21<br>(5.32) | 149<br>(10.30) | 49<br>(1.58) | 29<br>(1.04) | 33,824<br>(1.07) | 22<br>(0.08) | 728<br>(0.06) | 32,987<br>(6.15) | 31,059<br>(12.47) | 34,229<br>(2.07) |
| Hospitalization with pneumonia requiring intensive services or death | 29,905<br>(1.44) | 134<br>(4.91) | 22<br>(1.11) | N/A | 5<br>(1.27) | 38<br>(2.63) | 5<br>(0.16) | 3<br>(0.11) | 4,856<br>(0.02) | 24<br>(0.08) | 65<br>(0.01) | 7,226<br>(1.35) | 3,628<br>(1.46) | 7,368<br>(0.45) |
| (Outcome<br>proportion %) |  |  |  |  |  |  |  |  |  |  |  |  |  |  |
| Death | 11,407 | 335 | 43 | 406 | 1 | 43 | 5 | 4 | 965 | 24 | 75 | 2,603 | 1,354 | 3,513 |
| (Outcome<br>proportion %) | (0.55) | (12.27) | (2.17) | (1.07) | (0.25) | (2.97) | (0.16) | (0.14) | (0.03) | (0.08) | (0.01) | (0.48) | (0.54) | (0.21) |
| Age (% above<br>65) | 26.1 | 38.9 | 15.6 | 17.9 | 18.2 | 37.3 | 11.9 | 23.1 | 12.5 | 16.9 | 16.0 | 14.2 | 96.2 | 30.0 |
| Sex (% male) | 44.4 | 47.2 | 43.5 | 43.4 | 49.6 | 81.4 | 41.7 | 44.5 | 42.7 | 43.7 | 56.8 | 29.2 | 45.9 | 40.1 |
| Cancer (%) | 12.6 | 17.1 | 9.8 | 6.3 | 11.6 | 17.0 | 7.7 | 8.2 | 6.2 | 3.7 | 2.5 | 8.9 | 35.2 | 10.6 |
| COPD (%) | 10.2 | 9.3 | 4.9 | 2.5 | 6.3 | 20.5 | 2.7 | 3.1 | 2.7 | 2.7 | 0.5 | 19.8 | 26.6 | 7.6 |
| Diabetes (%) | 20.5 | 30.9 | 23.1 | 8.0 | 19.7 | 35.2 | 3.8 | 13.0 | 11.4 | 6.7 | 8.3 | 27.4 | 36.1 | 15.3 |
| Heart disease<br>(%) | 31.0 | 40.1 | 17.1 | 11.2 | 25.8 | 44.7 | 7.7 | 12.9 | 16.5 | 7.5 | 8.0 | 36.1 | 68.2 | 23.4 |
| Hypertension<br>(%) | 44.2 | 51.6 | 26.3 | 14.8 | 38.5 | 63.0 | 13.9 | 27.0 | 29.1 | 12.4 | 11.4 | 49.8 | 80.4 | 36.1 |
| Hyperlipidemia<br>(%) | 46.8 | 40.6 | 39.9 | 11.4 | 32.9 | 62.5 | 3.3 | 20.2 | 21.8 | 4.6 | 15.2 | 36.0 | 69.6 | 34.2 |
| Kidney disease<br>(%) | 18.7 | 31.2 | 17.0 | 11.0 | 24.3 | 32.4 | 7.6 | 6.2 | 9.0 | 1.2 | 5.1 | 23.4 | 35.5 | 14.9 |

In the databases used for external validation, the patient numbers ranged from 395 (TRDW) to 3,146,743 (CCAE). The datasets had varied outcome proportions ranging from 0.06-12.47 for hospital admission, 0.01-4.91 for intensive services, and 0.01-12.27 for fatality. Characteristics at baseline differed substantially between databases as can be seen in Table 2, with MDCR (a database representing retirees) containing a relatively old population of patients and a high number of comorbidities, and IPCI (a database representing general practice) showing a relatively low condition occurrence.

### Model specification

The data-driven models for hospitalization, intensive services, and fatality contained 521, 349, and 205 predictors respectively. The COVER-H, COVER-I, and COVER-F scores are presented in Figure 2. These models are accessible online.

**Fig 2.**
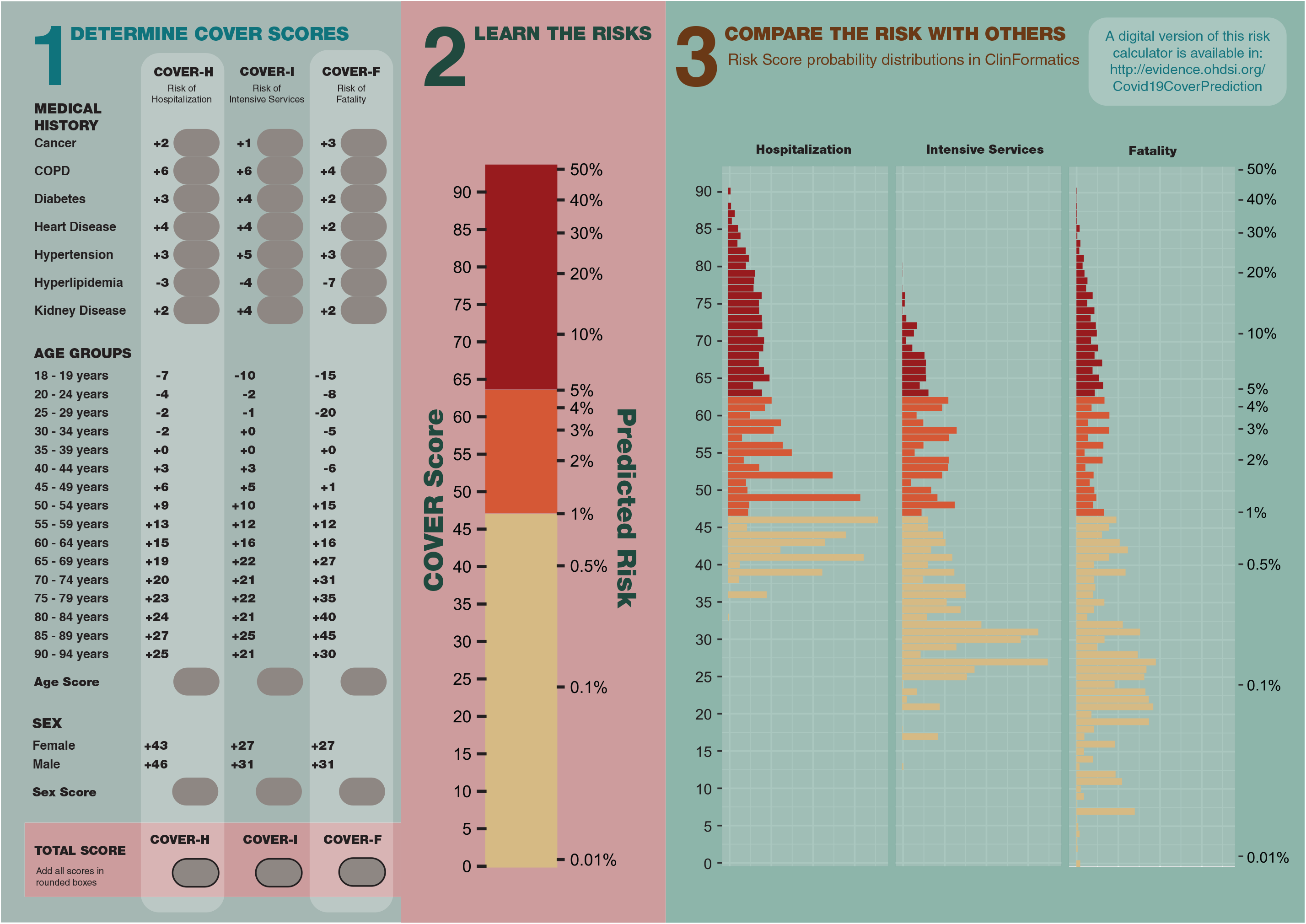

Figure 2 also provides a risk converter, which allows for easy conversion between the risk score and predicted risk of the outcomes^=^. The scores can be converted to a probability by applying the logistic function: 1/(1+exp((risk score-93)/10)). Furthermore, we provide a plot of the probability distribution for the three models from patients in ClinFormatics to demonstrate the expected regions the probabilities fall into. To calculate the COVER scores using Figure 2, a clinician needs to identify which predictors the patient has. The points for each of those predictors are then added to arrive at the total score. For example, if a 63-year-old female patient has diabetes and heart disease, then her risk score for hospital admission (COVER-H) is 43 (female sex) + 4 (heart disease) + 3 (diabetes) + 15 (age) = 65. The risk scores for intensive services (COVER-I) and fatality (COVER-F) are 51 and 47, respectively. Using the risk converter in Figure 2, a score of 65 corresponds to a risk of 6%. Scores of 51 and 47 correspond to 1.5% and 1%, respectively.

### Model performance

The internal validation performance for each model is presented in Table 3. The external validation of the COVER scores on the COVID-19 patients is shown in Table 4. Full validation results can be seen in Appendix 1B of the online supplement. Receiver operating characteristic and calibration plots are included in Appendix 1C of the online supplement.

**Table 3.** The results for internal validation in ClinFormatics

| Outcome | Predictors | No. Variables | AUC | AUPRC |
| --- | --- | --- | --- | --- |
| Hospitalization with pneumonia | Conditions/drugs<br>+ age/sex | 521 | 0.852 | 0.224 |
|  | Age/sex | 2 | 0.818 | 0.164 |
|  | COVER-H | 9 | 0.840 | 0.120 |
| Hospitalization with pneumonia requiring intensive services or death | Conditions/drugs<br>+ age/sex | 349 | 0.860 | 0.070 |
|  | Age/sex | 2 | 0.821 | 0.049 |
|  | COVER-I | 9 | 0.839 | 0.059 |
| Fatality | Conditions/drugs<br>+ age/sex | 205 | 0.926 | 0.069 |
|  | Age/sex | 2 | 0.909 | 0.037 |
|  | COVER-F | 9 | 0.896 | 0.039 |

**Table 4.** COVID-19 validation of the COVER scores on COVID-19 patients with a GP, ER, or OP visit in 2020 (*Confidence interval is not reported as the number of outcomes is larger than 1000)

| Outcome | Database | AUC (95% confidence interval) | AUPRC |
| --- | --- | --- | --- |
| Hospitalization<br>with pneumonia<br>(COVER-H) | HIRA | 0.806 (0.762-0.851) | 0.134 |
|  | SIDIAP | 0.748* | 0.072 |
|  | TRDW | 0.731 (0.611-0.851) | 0.132 |
|  | VA | 0.689 (0.649-0.729) | 0.179 |
| Hospitalization<br>with pneumonia<br>requiring<br>intensive services<br>or death<br>(COVER-I) | CUIMC | 0.734 (0.699-0.769) | 0.100 |
|  | HIRA | 0.910 (0.889-0.931) | 0.053 |
|  | VA | 0.763 (0.708-0.818) | 0.058 |
| Fatality<br>(COVER-F) | CUIMC | 0.820 (0.796-0.840) | 0.400 |
|  | HIRA | 0.898 (0.857-0.940) | 0.150 |
|  | SIDIAP | 0.895 (0.881-0.910) | 0.083 |
|  | VA | 0.717 (0.642-0.791) | 0.068 |

## Discussion

### Interpretation

We developed and externally validated models using large datasets of influenza patients to quantify a patient’s risk of developing severe or critical illness due to COVID-19. In the development data, the 9-predictor COVID-19 Estimated Risk (COVER) scores were a good trade- off between model complexity and performance, as the AUCs were generally close to the large data-driven models. The COVER scores achieved an AUC of 0.84 when predicting which patients will be hospitalized or require intensive services and an AUC of 0.9 when predicting which patients will die within 30 days. When validated on 1,985 COVID-19 patients in South Korea the COVER-H score performed well (AUC > 0.8), and COVER-I and COVER-F performed excellently (AUC ≥ 0.9). The model performed similarly well when applied to 37,950 COVID-19 Spanish patients (COVER-H: AUC 0.75) and excellent performance when predicting fatality (COVER-F: AUC 0.89). When applied to US patients, the COVER-I and COVER-F models achieved good AUCs of 0.73 and 0.82 in CUIMC, VA performed similarly with AUCs of 0.76 and 0.72 respectively. The VA also achieved 0.69 for COVER-H. A visual assessment of calibration plots across validations showed reasonable calibration in HIRA, SIDIAP, and VA. There was slight overestimation of risk amongst oldest and highest risk strata in SIDIAP, and to a lesser extent in HIRA. The calibration was poor in CUIMC, often underestimating risk, but this may be due to CUIMC containing mostly hospitalized COVID-19 patients, so the CUIMC cohort are experiencing more severe COVID-19 as due to the setting they are mostly hospital admitted patients when tested. The VA showed some miscalibration in the lowest and highest risk strata. The variable calibration results suggest that the models performance should be assessed and potentially recalibrated when used in a new context. We also performed sensitivity analyses using more sensitive COVID-19 definitions which also included patients with symptoms, or symptoms and influenza. The results did not show much deviation from the specific definition (Online supplement Appendix B).

These results showed that training in large historical influenza data was an effective strategy to develop models for COVID-19 patients. We also validated the age/sex and data-driven models on the COVID-19 patients and the age/sex models already appear to do well. This shows that age and sex are strong predictors of disease severity in COVID-19. Our results show that quantifying a symptomatic patient’s risk based on a small selection of comorbidities as well as age/sex gives improved model performance.

We identified one other model that addressed a similar problem setting. The COVID-19 Vulnerability Index built from a 5% sample of Medicare claims data from 2015-2016 using a proxy for COVID-19. The model predicts hospitalization due to pneumonia (except when caused by tuberculosis), influenza, acute bronchitis, or other specified upper respiratory infections^26^. The model achieved an AUC of 0.73, but has not been validated on a COVID-19 cohort. Several other models have been proposed to predict severity of COVID-19^27-29^, but these only consider patients already hospitalized.

### Limitations

Limitations of the study included being unable to develop a model on COVID-19 patient data due to the scarcity of databases that contain this information in sufficient numbers, however we were able to validate the models developed in COVID-19 and as such are confident the performance is transportable. In CUIMC, HIRA, SIDIAP, and VA COVID-19 databases we either reached or approached the threshold for reliable external validation of ∼100 patients who experience the outcome of interest^30,31^. The results of TRDW are promising, but might not be reliable due to the low number of outcomes. As larger COVID-19 databases become available, training a model using these data may highlight predictors of severity amongst uncommon influenza presentations, for example younger and healthier patients experiencing severe or critical illness.

The calibration in some of the COVID-19 validations could benefit from recalibration which can be performed by either recalibration in the large^32^ or logistic recalibration^33^. This suggests that calibration can be an issue in some locations and as such ideally the models will be tested and recalibrated in these locations before use.

Further limitations include misclassification of predictors, for example if disease is incorrectly recorded in a patient’s history, as well as in the cohorts through incorrect recording of influenza or COVID-19. We were unable to validate the COVER-H score in CUIMC as it mostly contained ER or hospitalized COVID-19 patients and the COVER-I score in SIDIAP due to a lack of information on intensive services in the database. A similar issue also meant we were not able to include some suspected disease predictors such as BMI/Obesity in the analysis due to the inconsistency with which these measures are collected and reported across the various databases included in the study.

We used a 30 risk window which has a limitation that if a patient experiences an outcome after the time window, this will be recorded as a non-event. This is unlikely for a hospital or intensive services admission, both of which tend to happen within 2 weeks of initial symptomatic presentation. Death has a higher probability of occurring outside this window but the likelihood of this is still small and so is unlikely to impact the performance evaluation significantly.

### Implications

The results show we were able to develop models that use a patient’s socio-demographics and medical history to predict their risk of becoming severely or critically ill when infected with COVID-19. To our knowledge, this is the first study that has been able to extensively externally validate prediction models on COVID-19 patients internationally. The strong performance in COVID-19 patients of the COVER scores can be used to identify patients who should be shielded from COVID-19. This can have multiple benefits; i) it can help reassure low risk people who may be psychologically impacted by the stress of the virus, and ii) it can help identify which people would be at increased risk of severe or critical outcomes and as such should continue to be shielded during the first stages of de-confinement.

### Conclusion

In this paper we developed and validated models that can predict which patients presenting with COVID-19 are at high risk of experiencing severe or critical illness. These models can be used to identify vulnerable patient populations that require shielding as they have the worst COVID-19 prognosis. This evidence can be particularly impactful as governments start to lift measures and could be used to aid strategic planning to help us protect the most vulnerable.

## Data Availability

Data is not publicly available due to patient privacy concerns.

## Acknowledgements

The authors would like to thank the OHDSI community for their contributions to the tools used for this analysis.

The authors appreciate healthcare professionals dedicated to treating COVID-19 patients in Korea, and the Ministry of Health and Welfare and the Health Insurance Review & Assessment Service of Korea for sharing invaluable national health insurance claims data in a prompt manner.

## Author contributions

All authors made substantial contributions to the conception or design of the work; JMR and RDW led the data analysis; all authors were involved in the analysis and interpretation of data for the work; all authors have contributed to the drafting and revising critically the manuscript for important intellectual content; all authors have given final approval and agree to be accountable for all aspects of the work.

## References

1. World Health Organization. Clinical management of severe acute respiratory infection (S ARI) when COVID-19 disease is suspected: interim guidance, 13 March 2020. Geneva: World Health Organization;2020.

2. Prieto-Alhambra D, Ballo E, Coma-Redon E, et al. Hospitalization and 30-day fatality in 121,263 COVID-19 outpatient cases. medRxiv. 2020:2020.2005.2004.20090050.

3. Anderson RM, Heesterbeek H, Klinkenberg D, Hollingsworth TD. How will country-based mitigation measures influence the course of the COVID-19 epidemic? The Lancet. 2020;395(10228):931–934.

4. Department of Health And Social Care. Coronavirus: action plan. In:2020.

5. Lee VJ, Chiew CJ, Khong WX. Interrupting transmission of COVID-19: lessons from containment efforts in Singapore. J Travel Med. 2020.

6. Wang CJ, Ng CY, Brook RH. Response to COVID-19 in Taiwan: Big Data Analytics, New Technology, and Proactive Testing. JAMA. 2020.

7. Saez M, Tobias A, Varga D, Barceló MA. Effectiveness of the measures to flatten the epidemic curve of COVID-19. The case of Spain. Sci Total Environ. 2020;727:138761.

8. Thunström L, Newbold SC, Finnoff D, Ashworth M, Shogren JF. The benefits and costs of using social distancing to flatten the curve for COVID-19. Journal of Benefit-Cost Analysis. 2020:1–27.

9. Markel H, Lipman HB, Navarro JA, et al. Nonpharmaceutical Interventions Implemented by US Cities During the 1918-1919 Influenza Pandemic. JAMA. 2007;298(6):644–654.

10. New York State. A guide to reopening New York & building back better. 2020.

11. UK Government. Our plan to rebuild: The UK Government’s COVID-19 recovery strategy. 2020.

12. Rijksoverheid. Factsheet versoepelen maatregelen corona. 2020.

13. Burn E, You SC, Sena A, et al. An international characterisation of patients hospitalised with COVID-19 and a comparison with those previously hospitalised with influenza. medRxiv. 2020:2020.2004.2022.20074336.

14. Scally G, Jacobson B, Abbasi K. The UK’s public health response to covid-19. BMJ. 2020;369:m1932.

15. World Health Organization. Coronavirus disease 2019 (COVID-19) Situation report - 51 2020, 11 March 2020. World Health Organization;2020.

16. Wynants L, Van Calster B, Bonten MMJ, et al. Prediction models for diagnosis and prognosis of covid-19 infection: systematic review and critical appraisal. BMJ. 2020;369:m1328.

17. Miyashita N, Matsushima T, Oka M, Japanese Respiratory S. The JRS guidelines for the management of community-acquired pneumonia in adults: an update and new recommendations. Intern Med. 2006;45(7):419–428.

18. Fan G, Tu C, Zhou F, et al. Comparison of severity scores for COVID-19 patients with pneumonia: a retrospective study. Eur Respir J. 2020.

19. Reps JM, Schuemie MJ, Suchard MA, Ryan PB, Rijnbeek PR. Design and implementation of a standardized framework to generate and evaluate patient-level prediction models using observational healthcare data. J Am Med Inform Assoc. 2018;25(8):969–975.

20. Reps JM, Williams RD, You SC, et al. Feasibility and evaluation of a large-scale external validation approach for patient-level prediction in an international data network: validation of models predicting stroke in female patients newly diagnosed with atrial fibrillation. BMC Med Res Methodol. 2020;20(1):102–102.

21. Overhage JM, Ryan PB, Reich CG, Hartzema AG, Stang PE. Validation of a common data model for active safety surveillance research. J Am Med Inform Assoc. 2012;19(1):54–60.

22. Suchard MA, Simpson SE, Zorych I, Ryan P, Madigan D. Massive parallelization of serial inference algorithms for a complex generalized linear model. ACM Transactions on Modeling and Computer Simulation (TOMACS). 2013;23(1):1–17.

23. Steyerberg EW, Vergouwe Y. Towards better clinical prediction models: seven steps for development and an ABCD for validation. Eur Heart J. 2014;35(29):1925–1931.

24. Moons KG, Altman DG, Reitsma JB, et al. Transparent Reporting of a multivariable prediction model for Individual Prognosis or Diagnosis (TRIPOD): explanation and elaboration. Ann Intern Med. 2015;162(1):W1–73.

25. Observational Health Data Sciences and Informatics. The Book of OHDSI. 2019.

26. DeCaprio D, Gartner J, Burgess T, Kothari S, Sayed S. Building a COVID-19 Vulnerability Index. arXiv preprint 200307347. 2020.

27. Liang W, Liang H, Ou L, et al. Development and Validation of a Clinical Risk Score to Predict the Occurrence of Critical Illness in Hospitalized Patients With COVID-19. JAMA Internal Medicine. 2020.

28. Xiao L-s, Zhang W-F, Gong M, et al. Development and Validation of the HNC-LL Score for Predicting the Severity of Coronavirus Disease 2019. Available at SSRN 3572843. 2020.

29. Ji D, Zhang D, Xu J, et al. Prediction for Progression Risk in Patients with COVID-19 Pneumonia: the CALL Score. Clin Infect Dis. 2020.

30. Vergouwe Y, Steyerberg EW, Eijkemans MJ, Habbema JDF. Substantial effective sample sizes were required for external validation studies of predictive logistic regression models. J Clin Epidemiol. 2005;58(5):475–483.

31. Collins GS, Ogundimu EO, Altman DG. Sample size considerations for the external validation of a multivariable prognostic model: a resampling study. Stat Med. 2016;35(2):214–226.

32. Toll DB, Janssen KJ, Vergouwe Y, Moons KG. Validation, updating and impact of clinical prediction rules: a review. J Clin Epidemiol. 2008;61(11):1085–1094.

33. Steyerberg EW, Borsboom GJ, van Houwelingen HC, Eijkemans MJ, Habbema JD. Validation and updating of predictive logistic regression models: a study on sample size and shrinkage. Stat Med. 2004;23(16):2567–2586.

